# Outcomes of Patients with Subacute/Chronic DeBakey Type I Aortic Dissection: A Multicenter Study

**DOI:** 10.64898/2026.01.05.26343491

**Authors:** Hao Chen, Hao Han, Xiaomeng Wang, Shi Yue, Youjin Li, Haiping Yang, Hongli Li, Gang Liu, Minjia Zhu, Jian Huang, Qingwu Zhao, Jihong Liu, Haibo Wu, Junming Zhu, Wenjian Jiang, Yuan Xue, Haiyang Li, Hongjia Zhang

## Abstract

**BACKGROUND:** The optimal surgical strategy for subacute and chronic DeBakey type I aortic dissection remains controversial. This multicenter study compared early outcomes and long-term survival between total arch replacement (TAR) with a frozen elephant trunk (FET) and conventional non-FET repair.

**METHODS:** We retrospectively analyzed 452 patients who underwent surgery for subacute or chronic DeBakey type I aortic dissection at seven high-volume aortic centers between 2015 and 2023. Of these, 320 patients received TAR+FET and 132 underwent non-FET procedures. Perioperative outcomes and long-term survival were assessed using inverse probability of treatment weighting (IPTW), Kaplan–Meier survival curves, and landmark analysis.

**RESULTS:** After IPTW adjustment, thirty-day mortality was higher in the FET group than in the non-FET group (6.2% vs. 2.0%, P=0.044). However, there was no significant difference in overall survival between the two groups (log-rank P = 0.469). The landmark analysis showed no difference in all-cause mortality within the first six years, but mortality was significantly lower in the FET group thereafter (1.8% vs. 12.5%, P=0.007).

**CONCLUSIONS:** In patients with subacute or chronic DeBakey type I dissection, TAR+FET is a feasible surgical method associated with increased early mortality, but potential late survival benefit compared with non-FET approaches. These findings warrant confirmation in prospective studies with extended follow-up.

## INTRODUCTION

DeBakey type I aortic dissection is a life-threatening form of aortic disease that requires immediate surgical intervention because of its high early mortality rate^1^. According to the 2024 EACTS/STS Guidelines, aortic dissection is categorized into four temporal phases: acute (1–14 days), subacute (15–90 days), and chronic (>90 days)^2^. Patients with acute DeBakey type I dissection rarely survive to the subacute or chronic stage due to rapid hemodynamic deterioration and prompt surgical treatment^3^. However, delayed diagnosis and uneven distribution of aortic centers—particularly in less developed regions of China—may result in a subset of patients presenting in the subacute or chronic stage after referral to high-volume centers^3^.

Existing studies on subacute or chronic DeBakey type I dissection are limited and largely descriptive^4,5^. Previous studies have primarily focused on comparing clinical characteristics and outcomes between acute and subacute/chronic cases. However, these investigations have not sufficiently explored the impact of various surgical strategies on long-term prognosis^3,6^. As a result, the best approach for surgical repair in this group is still debated.

Conservative repair strategies, such as ascending or hemiarch replacement, are commonly selected to minimize surgical trauma and perioperative risk^6,7^. However, residual dissection in the distal aorta may continue to dilate, predisposing patients to rupture or reintervention during follow-up. In contrast, total arch replacement (TAR) combined with frozen elephant trunk (FET) has emerged as an integrated approach allowing both open and endovascular treatment of the distal arch and proximal descending aorta, promoting false lumen thrombosis and favorable aortic remodeling^8,9^. Although some single-center studies show promising short-term results for TAR+FET in chronic dissection, there is a lack of strong multicenter evidence comparing TAR+FET to less extensive non-FET repairs in subacute or chronic DeBakey type I dissection.

Therefore, we conducted a multicenter, retrospective cohort study to compare early outcomes and long-term survival between TAR+FET and non-FET repair in patients with subacute or chronic DeBakey type I aortic dissection.

## PATIENTS AND METHODS

### PATIENTS

**Supplemental Figure 1** shows the study design. Between February 2015 and September 2023, 3,035 patients with Stanford type A aortic dissection underwent surgical treatment at seven tertiary aortic centers in China: Beijing Anzhen Hospital, People’s Hospital of Ningxia Hui Autonomous Region, The First Hospital of Hebei Medical University, The First Affiliated Hospital of Xiamen University, Guangdong Provincial Hospital of Traditional Chinese Medicine, Affiliated Zhongshan Hospital of Dalian University, and The Second Affiliated Hospital of Dalian Medical University. Among them, 452 patients were diagnosed with subacute or chronic DeBakey type I aortic dissection and met the inclusion criteria for this retrospective study.

Patients with acute dissection (<15 days from symptom onset) (n = 2492), and DeBakey type II dissection (n = 91) were excluded. The study was approved by the Institutional Review Board of Beijing Anzhen Hospital (KS2023090, approved December 8, 2023) and registered in the Chinese Clinical Trial Registry (ChiCTR1900022637). Because anonymized data were used for analysis, informed consent was waived.

Each participating center contributed data from its prospectively maintained aortic database, which were retrospectively pooled and verified by the coordinating center (Beijing Anzhen Hospital). Data integrity and variable definitions were standardized across centers before analysis. Follow-up information was obtained through review of outpatient records and structured telephone interviews. Long-term survival data were verified through hospital databases or official death registries when available.

### SURGICAL APPROACH

The surgical technique followed the reported approach for aortic dissection at our institution^8^, all participating centers adhered to standardized surgical principles for aortic arch repair, ensuring procedural consistency across institutions. All operations were performed via median sternotomy under cardiopulmonary bypass (CPB) with unilateral or bilateral selective cerebral perfusion (SCP). Arterial inflow was established using either the right axillary artery, femoral artery, or a dual arterial cannulation (DAC) strategy, selected according to patient anatomy and surgeon preference. In general, moderate hypothermia (nasopharyngeal temperature 22–30 °C) was applied. The DAC approach was preferred in patients with extensive dissection, multivessel malperfusion, or true lumen compression to ensure stable systemic perfusion, based on our experience and previous report^10^. USCP was routinely utilized, with BSCP being considered as a supplemental strategy when inadequate perfusion flow is encountered.

For total arch replacement with FET, a stent graft (MicroPort Medical Co., Shanghai, China) was deployed into the true lumen of the descending aorta, and the distal aortic arch was reconstructed using a four-branch prosthetic graft (Boston Scientific Inc., USA). In patients with false lumen perfusion, the FET procedure is generally avoided to reduce the risk of distal organ malperfusion. Distal anastomosis was performed between the left subclavian and left carotid arteries under circulatory arrest. Perfusion was resumed through the prosthetic graft after completion of the distal anastomosis.

In the non-FET group, patients underwent ascending aortic replacement, hemiarch replacement, or total arch replacement without stent graft implantation, depending on the extent of dissection and surgeon judgment.

### CLINICAL ENDPOINTS

The primary endpoints were thirty-day mortality and long-term all-cause mortality. Secondary endpoints included ICU retention time, ventilation time, stroke, paraplegia, acute kidney injury requiring dialysis, reoperation for bleeding and secondary intervention.

### DEFINITIONS

The diagnosis of subacute or chronic DeBakey type I aortic dissection was based on the following three criteria. First, the time from symptom onset to hospital admission was more than 14 days^2^. Second, computed tomography scans revealed the following typical signs: non-wavy, fixed, and stiff intimal flap, significant enlargement of the false lumen, and/or thrombus presence in the false lumen. Third, subacute or chronic DeBakey type I aortic dissection was confirmed by intraoperative identification of thickened and fibrotic changes in the intimal flap or by postoperative pathological results. Thirty-day mortality was defined as death from any cause occurring within 30 days after surgery, regardless of the patient’s location (whether in-hospital or after discharge). Stroke was defined as postoperative cerebral infarction or cerebral hemorrhage. Stroke could be accompanied by obvious or no neurological deficit symptoms, with relevant cerebral lesions confirmed by computed tomography and/or magnetic resonance imaging. Permanent neurologic deficit was defined as the impairment of neurologic function such as loss or indistinctness of speech, a change in the state of consciousness, or the presence of a motor deficit, which was caused by a disruption in the cerebral blood supply and persisted even after the patient’s discharge from the hospital^11^. Secondary intervention was defined as open reoperation of aorta or thoracic endovascular aortic repair (TEVAR).

### STATISTICAL ANALYSIS

Continuous variables were expressed as mean ± standard deviation or median (interquartile range). Categorical variables are presented as frequencies and percent. The Student’s t-test was used to compare normally distributed continuous variables, while the Mann-Whitney U test was applied for non-normally distributed continuous variables. The chi-square test and Fisher’s exact test were used for categorical variable comparisons. Two-tailed P < 0.05 was considered statistically significant.

The postoperative survival curve for all discharged patients was constructed using the Kaplan-Meier method, and group differences were assessed using log-rank tests. Landmark analyses were performed to assess survival rates by dividing the 9-year follow-up period into the first 6 years and the subsequent 3 years. Hazard ratios and 95% confidence intervals for 9-year survival rates were calculated using Cox proportional hazards regression models.

To minimize selection bias, inverse probability of treatment weighting (IPTW) was applied based on a propensity score including age, sex, body mass index (BMI), time of onset, baseline creatinine level, smoking, diabetes, hypertension, history of cerebral infarction, history of cardiovascular disease, history of aortic surgery, asymptomatic, maximum ascending aortic diameter, left ventricular ejection fraction (LVEF), aortic regurgitation, aortic root replacement, concomitant coronary artery bypass graft (CABG), concomitant mitral valve replacement (MVR), and concomitant Tricuspid valve repair (TVr). All analyses were conducted using R software (version 4.4.2).

## RESULTS

### PREOPERATIVE DATA

Preoperative characteristics of the unweighted and weighted cohorts are summarized in **Table 1 and Supplementary Table 1**. After IPTW adjustment, baseline variables were well balanced between the two groups (**Supplemental Figure 2**). The median age of the entire cohort was 51 years (IQR 41-60). Male patients constituted 69.47% of the cohort, with a significantly higher proportion in the FET group than in the non-FET group (74.69% vs 56.82%, P < 0.001). Hypertension was present in 69.91% of the patients (n = 316), and Marfan syndrome was identified in 1.6% (n = 7). Smoking was less prevalent in the non-FET group than in the FET group (27.27% vs. 45.94%, P < 0.001). In addition, more patients in the FET group had a prior history of aortic surgery (11.88% vs. 3.79%, P = 0.013). The median interval from symptom onset to surgery was 30 days (IQR 20–99). A total of 117 patients (39.16%) were asymptomatic at presentation. Among symptomatic patients, the most common manifestations were sudden pain (42.7%, n = 193), chest tightness (19.03%, n = 86), and diaphoresis (8.41%, n = 38) (Supplementary Table 1).

**Table 1.**
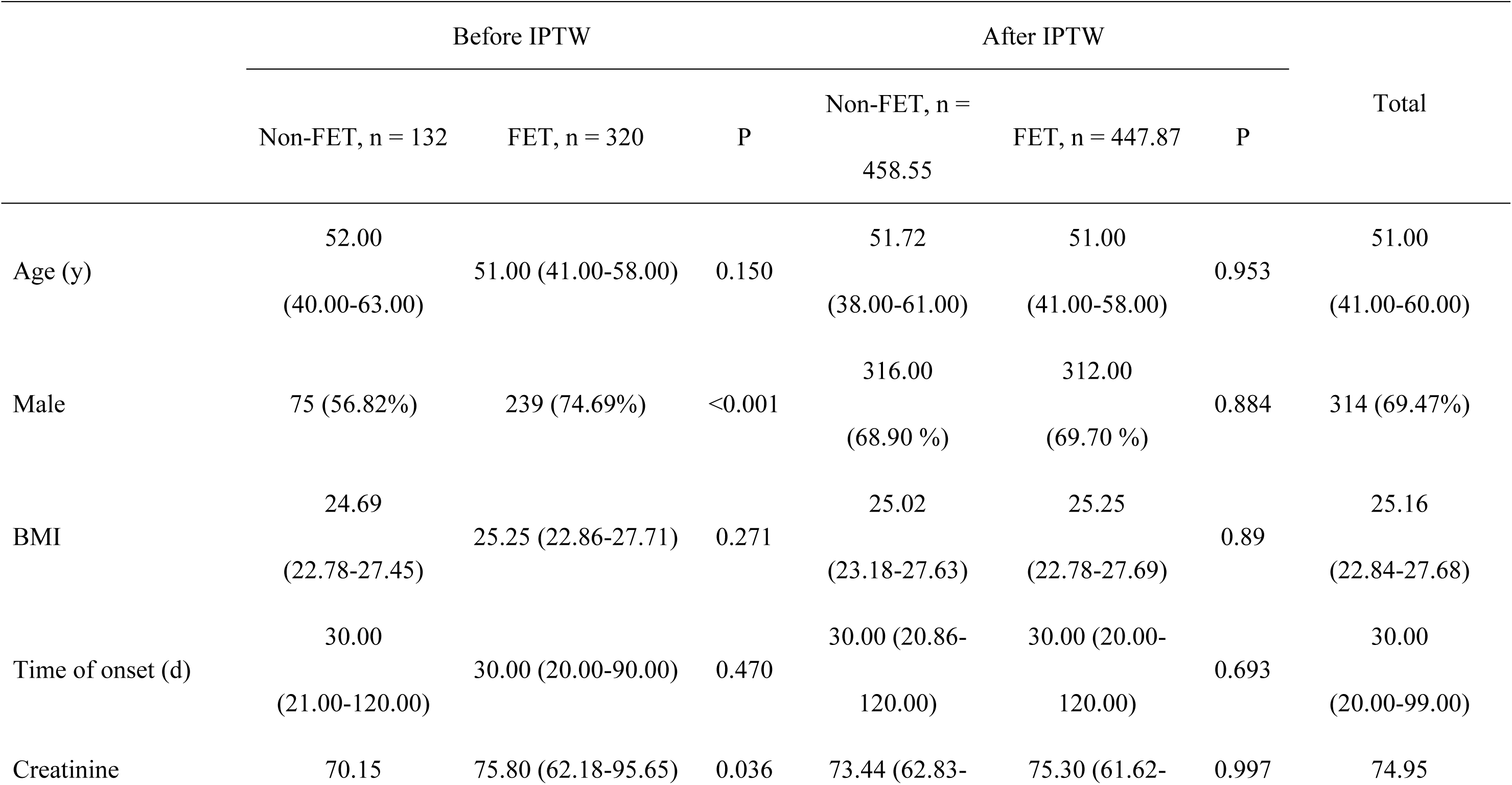
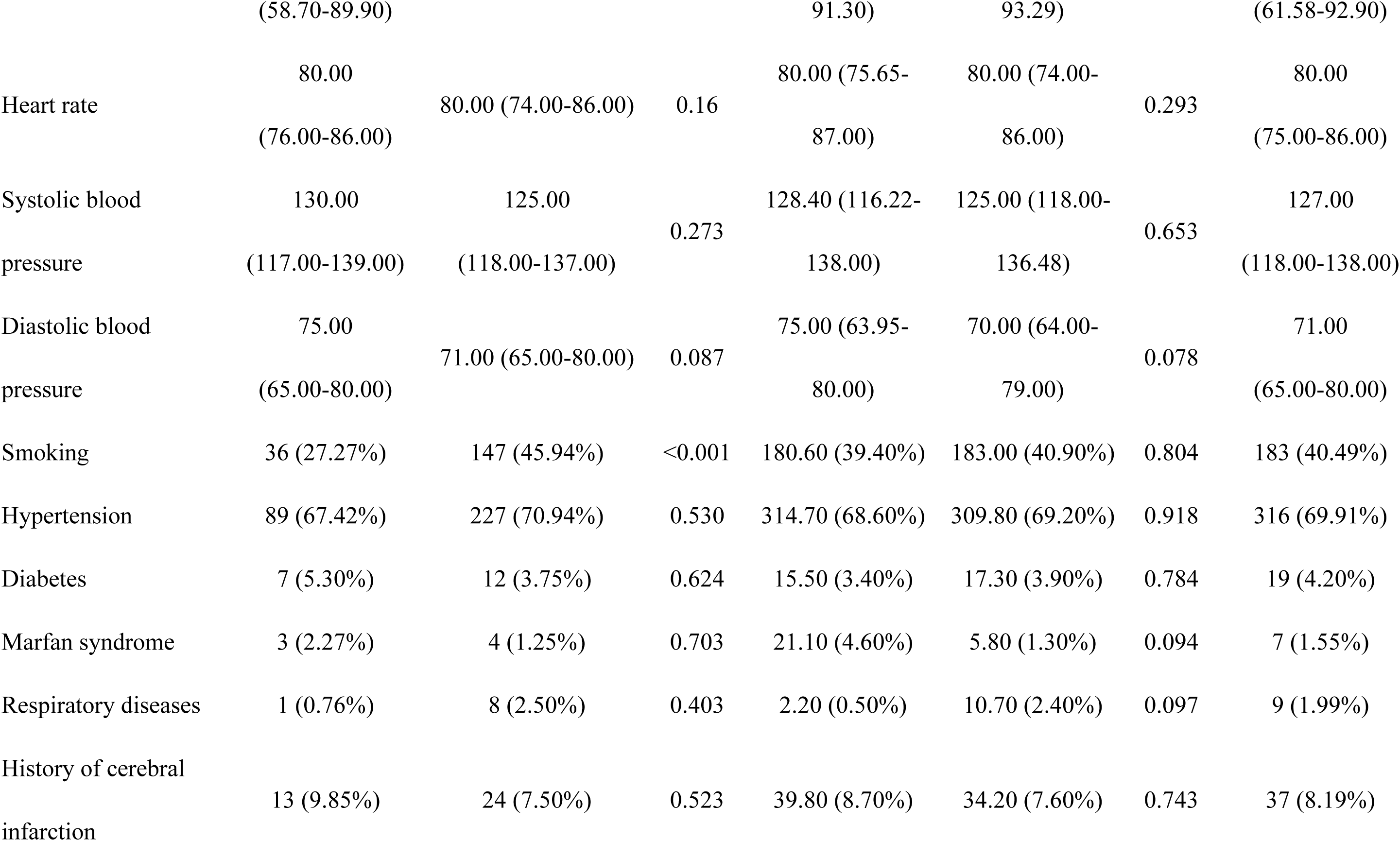
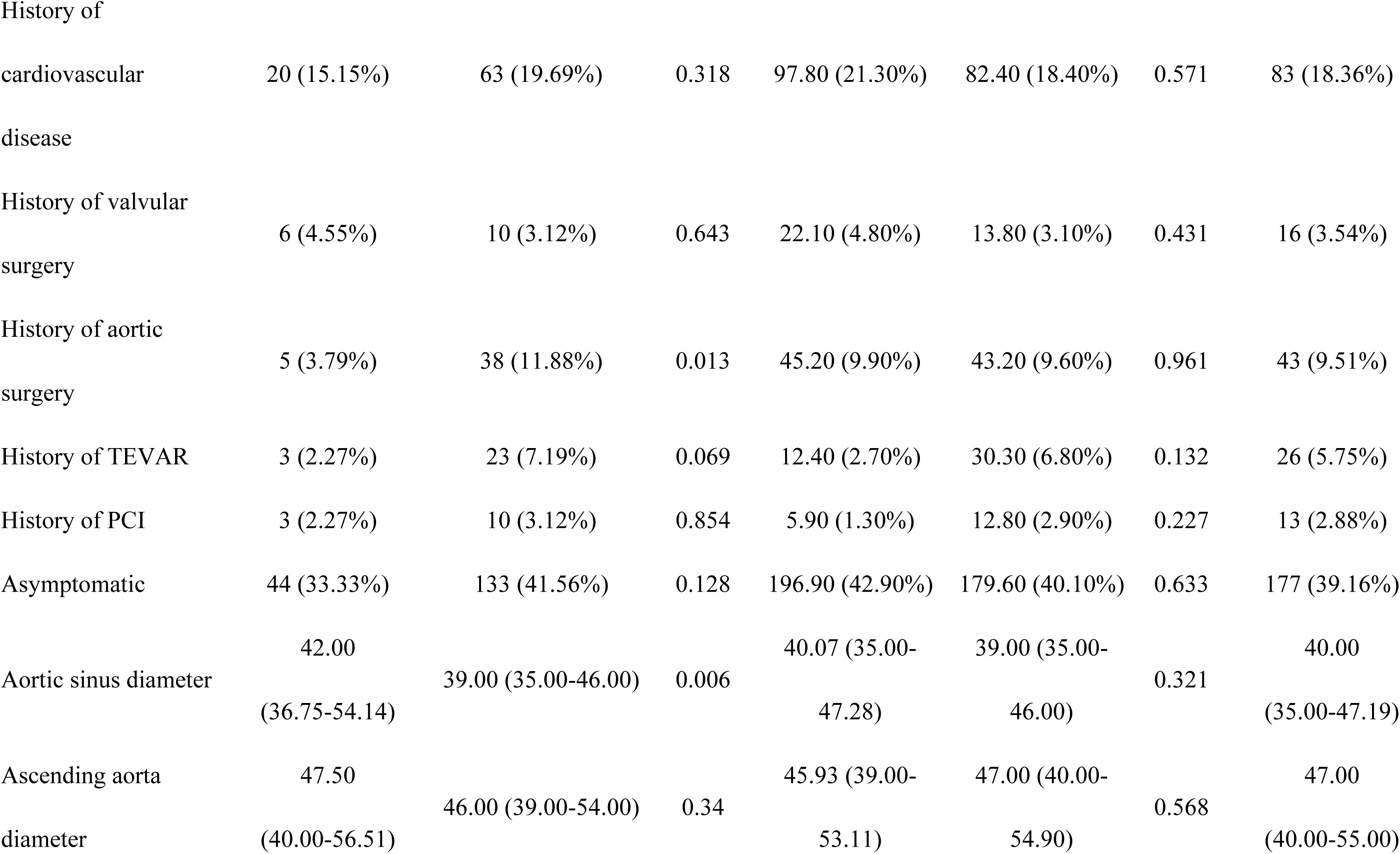
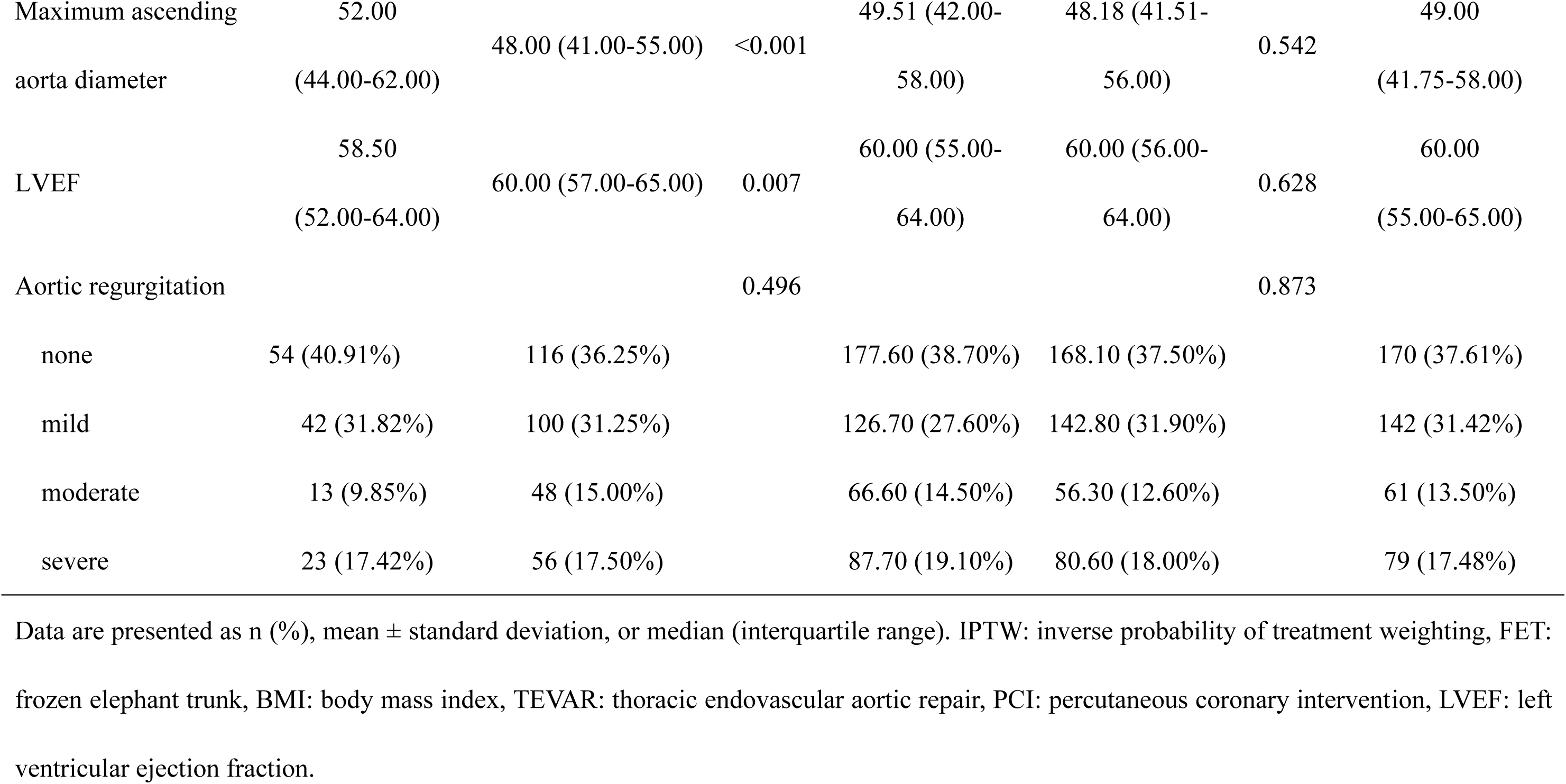
Preoperative characteristics of the total cohort and IPTW cohort.

Compared with the non-FET group, the FET group had smaller aortic sinus diameters [39.00 mm (IQR 35.00-46.00) vs. 42.00 mm (IQR 36.75-54.14), P = 0.006] and smaller maximum ascending aortic diameters [48.00 mm (IQR 41.00-55.00) vs. 52.00 mm (IQR 44.00-62.00), P < 0.001]. The LVEF was also significantly higher in the FET group [60.00% (IQR 57.00-65.00) vs. 58.50% (IQR 52.00-64.00), P = 0.007].

### INTRAOPERATIVE DATA

**Table 2** summarizes the intraoperative findings. In the non-FET group, 73 patients (55.30%) underwent isolated ascending aortic replacement, 57 (43.18%) underwent the Bentall procedure, and 2 (1.52%) received the Wheat procedure; none received the David procedure. In the FET group, 195 patients (60.94%) underwent isolated ascending aortic replacement, 122 (38.13%) underwent the Bentall procedure, 1 (0.31%) underwent the Wheat procedure, and 2 (0.62%) received the David procedure. Regarding arch interventions, 32 patients (24.24%) in the non-FET group had no arch intervention, 75 (56.82%) underwent hemiarch replacement, and 25 (18.94%) received total arch replacement. By design, all patients in the FET group underwent total arch replacement as part of the FET procedure.

**Table 2.**
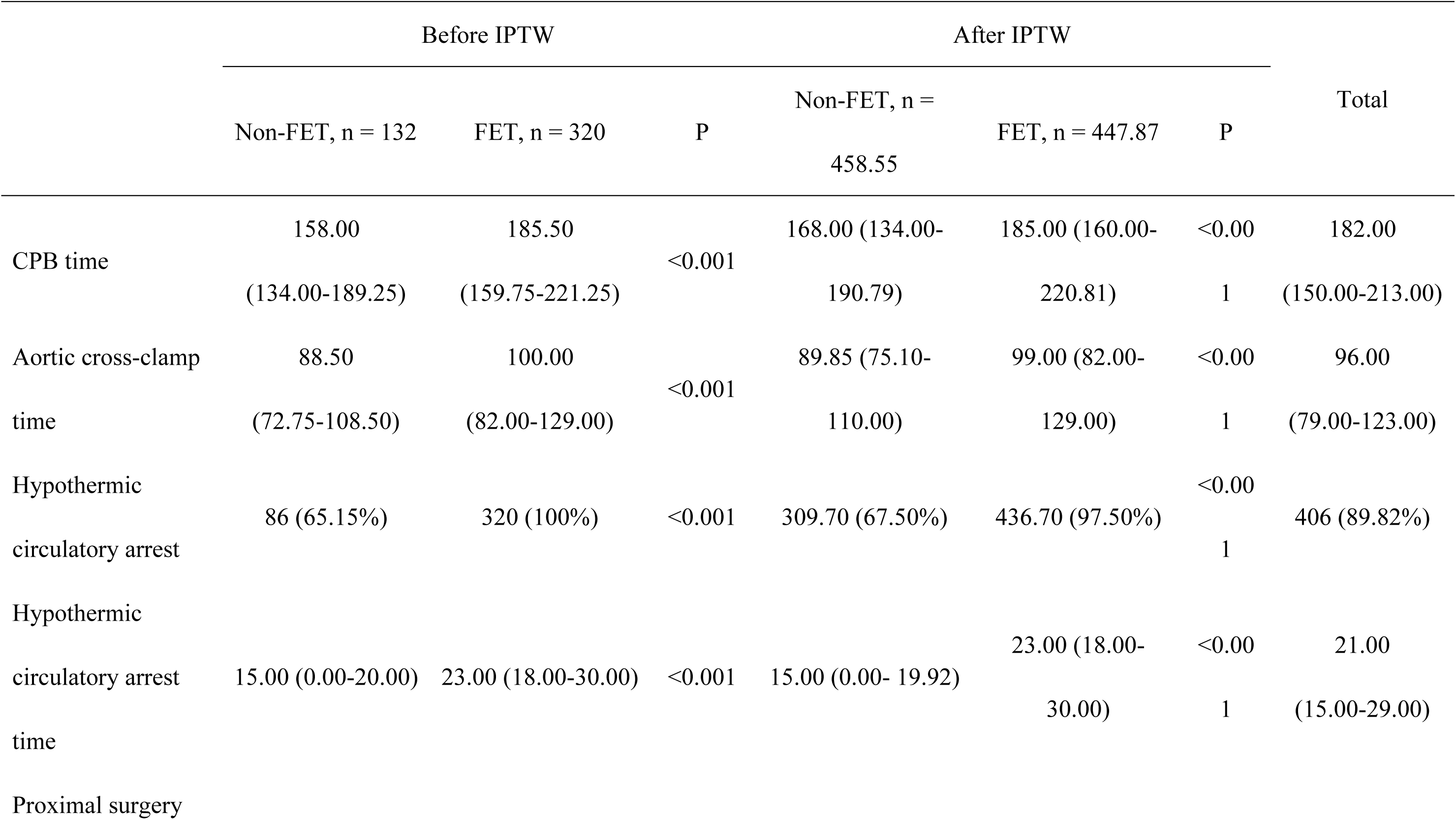
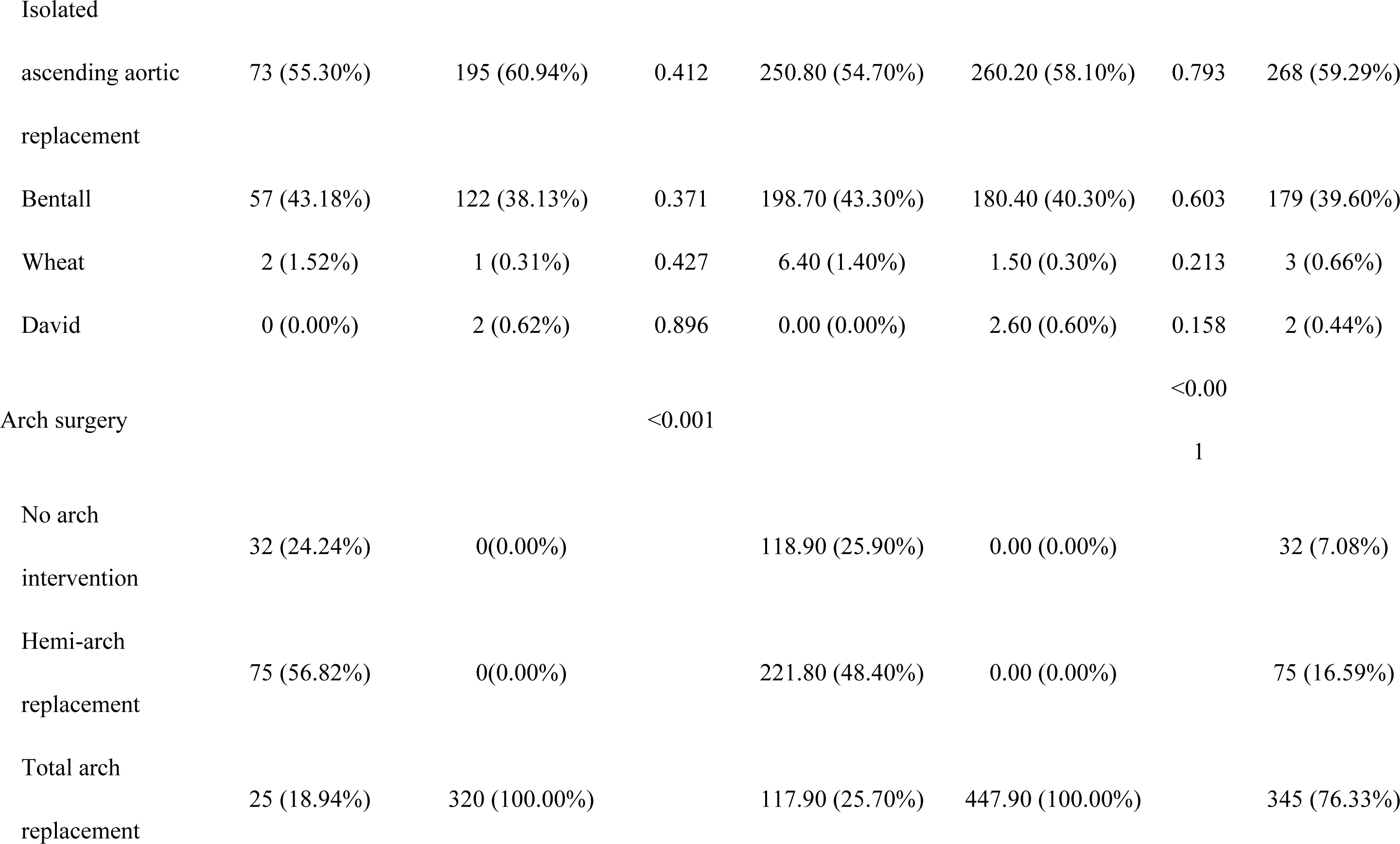
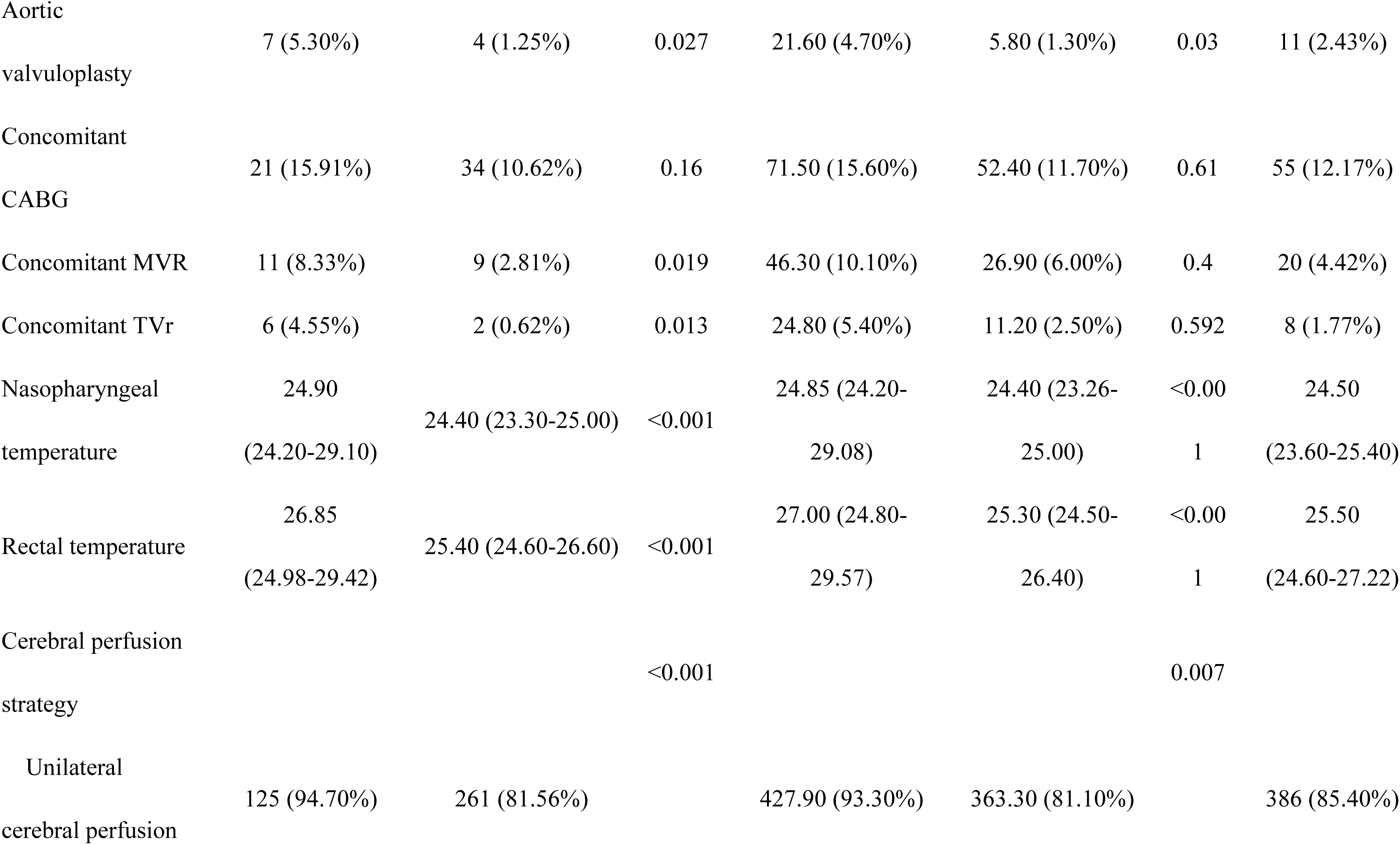
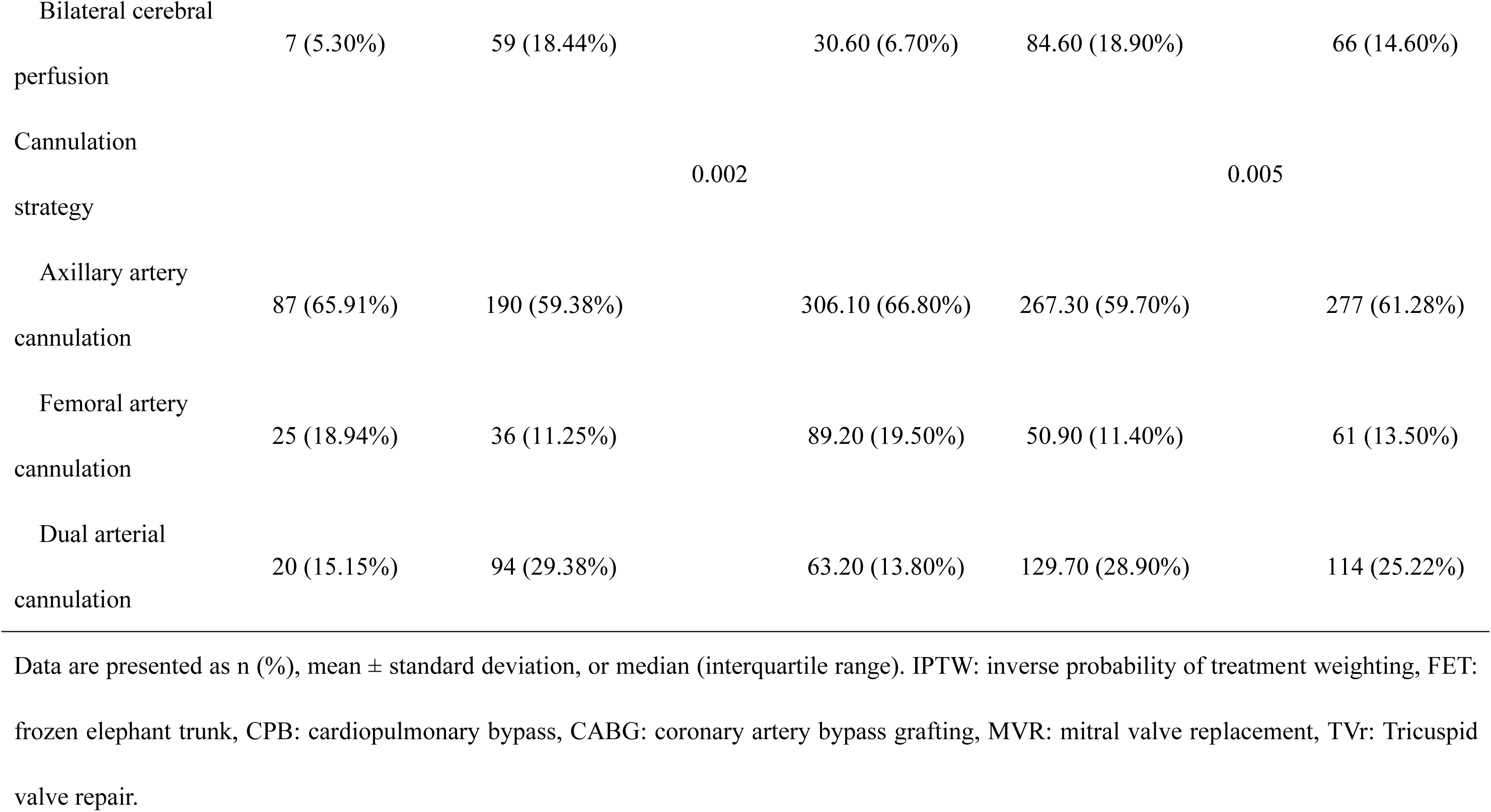
Intraoperative data of the total cohort and IPTW cohort.

Compared with the non-FET group, the FET group more frequently received bilateral selective cerebral perfusion (BSCP, 18.44% vs 5.30%, P < 0.001) and required longer cardiopulmonary bypass [185.50 min (IQR 159.75-221.25) vs 158.00 min (IQR 134.00-189.25), P < 0.001] and aortic cross-clamp times [100.00 min (IQR 82.00-129.00) vs 88.50 min (IQR 72.75-108.50), P < 0.001]. Intraoperative temperatures were significantly lower in the FET group, with nasopharyngeal temperature [24.40°C (IQR 23.30-25.00) vs 24.90°C (IQR 24.20-29.10), P < 0.001] and rectal temperature [25.40°C (IQR 24.60–26.60) vs 26.85°C (IQR 24.98–29.42), P < 0.001]. Fewer patients in the FET group underwent aortic valvuloplasty (1.25% vs 5.30%, P = 0.027), concomitant mitral valve replacement (2.81% vs 8.33%, P = 0.019), or tricuspid valve repair (0.62% vs 4.55%, P = 0.013).

After IPTW adjustment, significant differences persisted in CPB time, aortic cross-clamp time, hypothermic circulatory arrest, and aortic valvuloplasty, whereas differences in concomitant MVR and TVr were no longer significant.

### EARLY OUTCOMES

The overall 30-day mortality was 5.09% (n = 23), with 18 patients (5.6%) in the FET group and 5 patients (3.8%) in the non-FET group (P = 0.245) (**Figure 1**). Postoperative stroke occurred in 20 patients (4.42%), and paraplegia was observed in 8 (1.77%). Acute kidney injury requiring dialysis developed in 24 patients (5.31%). No significant differences were observed between groups for these outcomes before IPTW adjustment.

**Figure 1.**
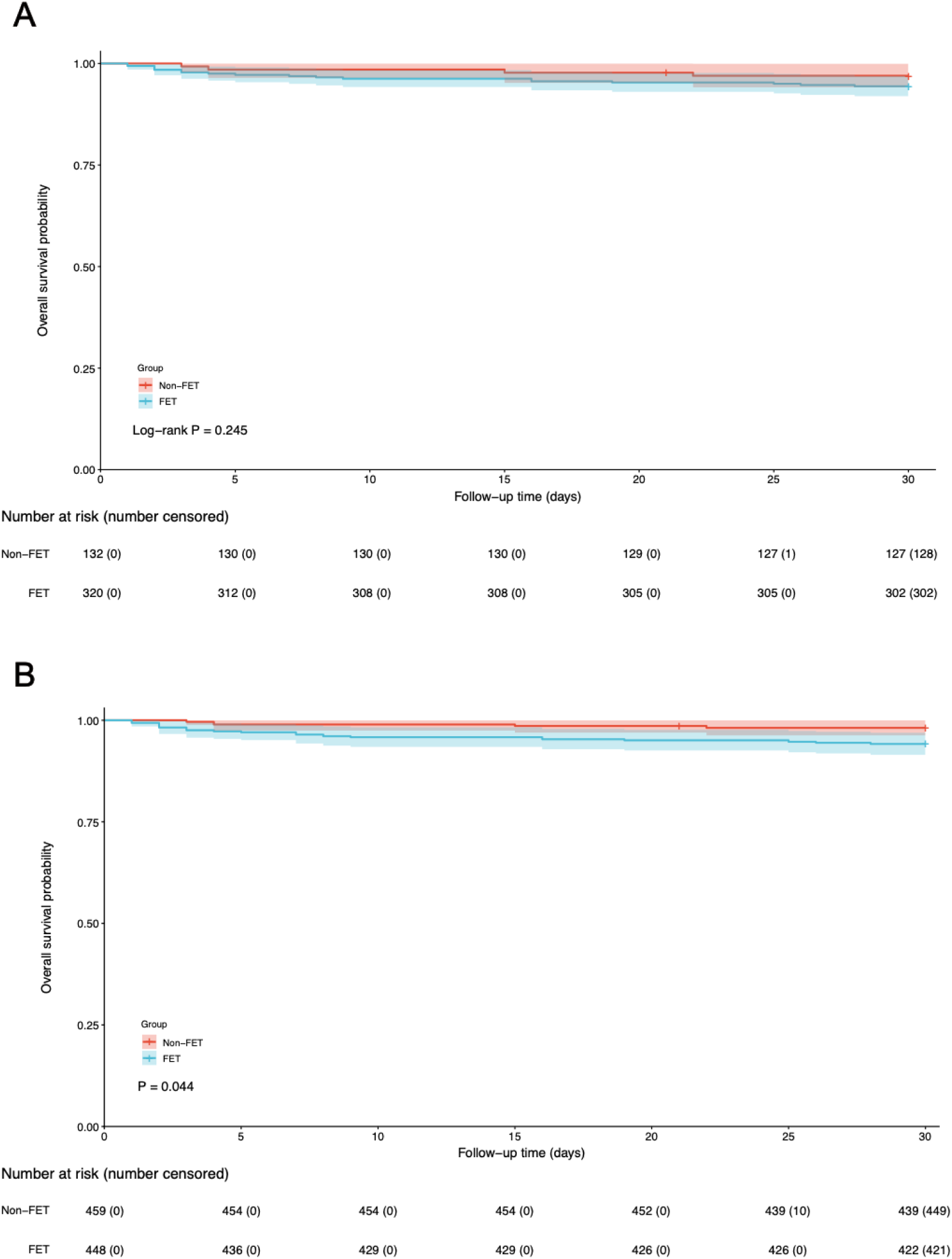
30-day Kaplan–Meier curve for survival before and after IPTW. A) Before IPTW, the 30-day survival rates were 94.37% for the FET group and 96.21% for the non-FET group (P = 0.25). B) After IPTW, the 30-day survival rates were 93.75% for the FET group and 97.82% for the non-FET group (P = 0.023). IPTW, inverse probability of treatment weighting, FET: frozen elephant trunk.

After IPTW adjustment, 30-day mortality was higher in the FET group than in the non-FET group (6.2% vs 2.0%, P = 0.044) (**Figure 1**). Permanent neurological injury also occurred more frequently in the FET group (2.0% vs 0%, P = 0.008). Furthermore, the incidence of postoperative pulmonary infection was significantly higher among FET patients (11.4% vs 5.2%, P = 0.036). No significant between-group differences were found for other postoperative complications after weighting (**Table 3**).

**Table 3.**
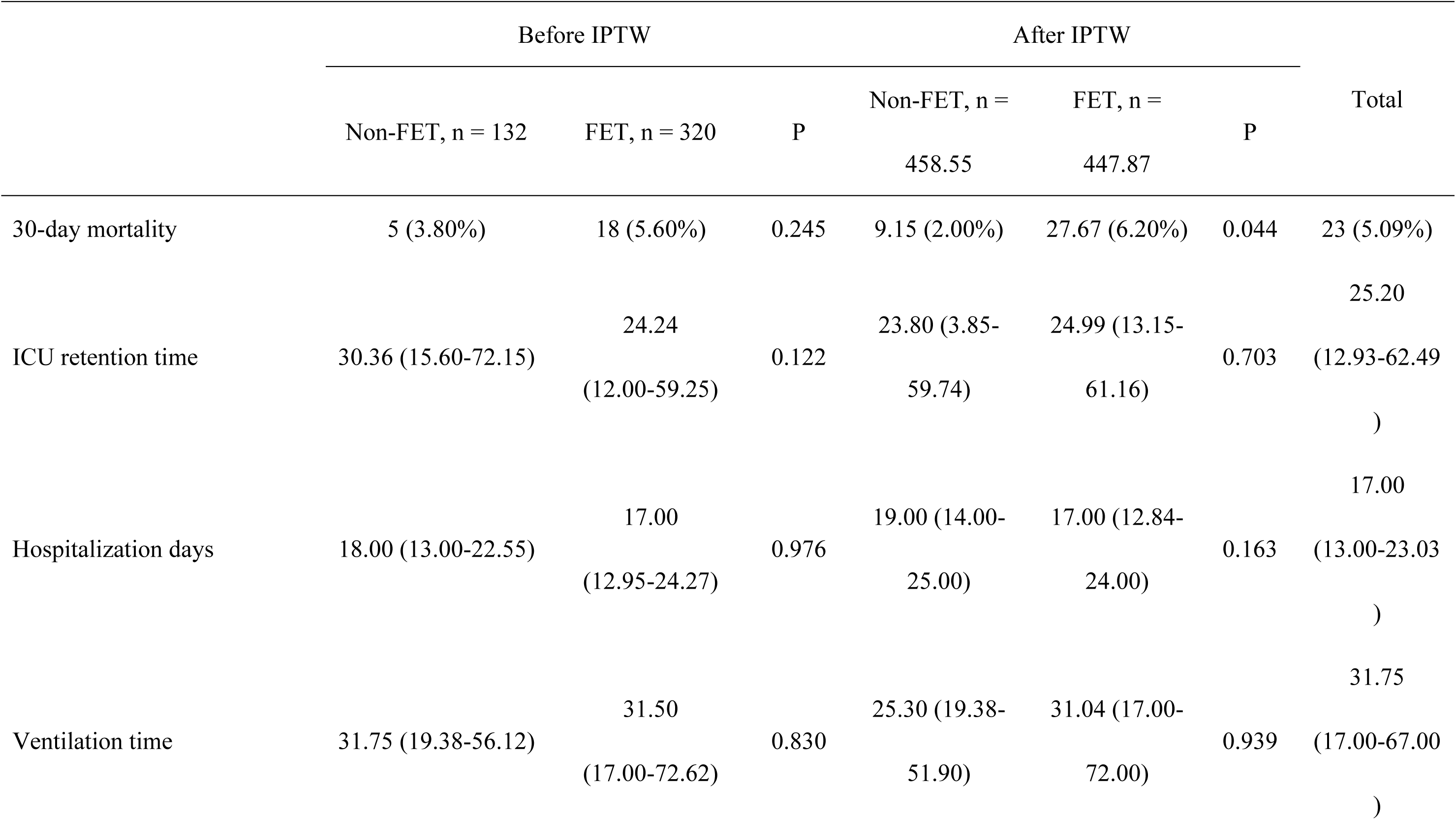
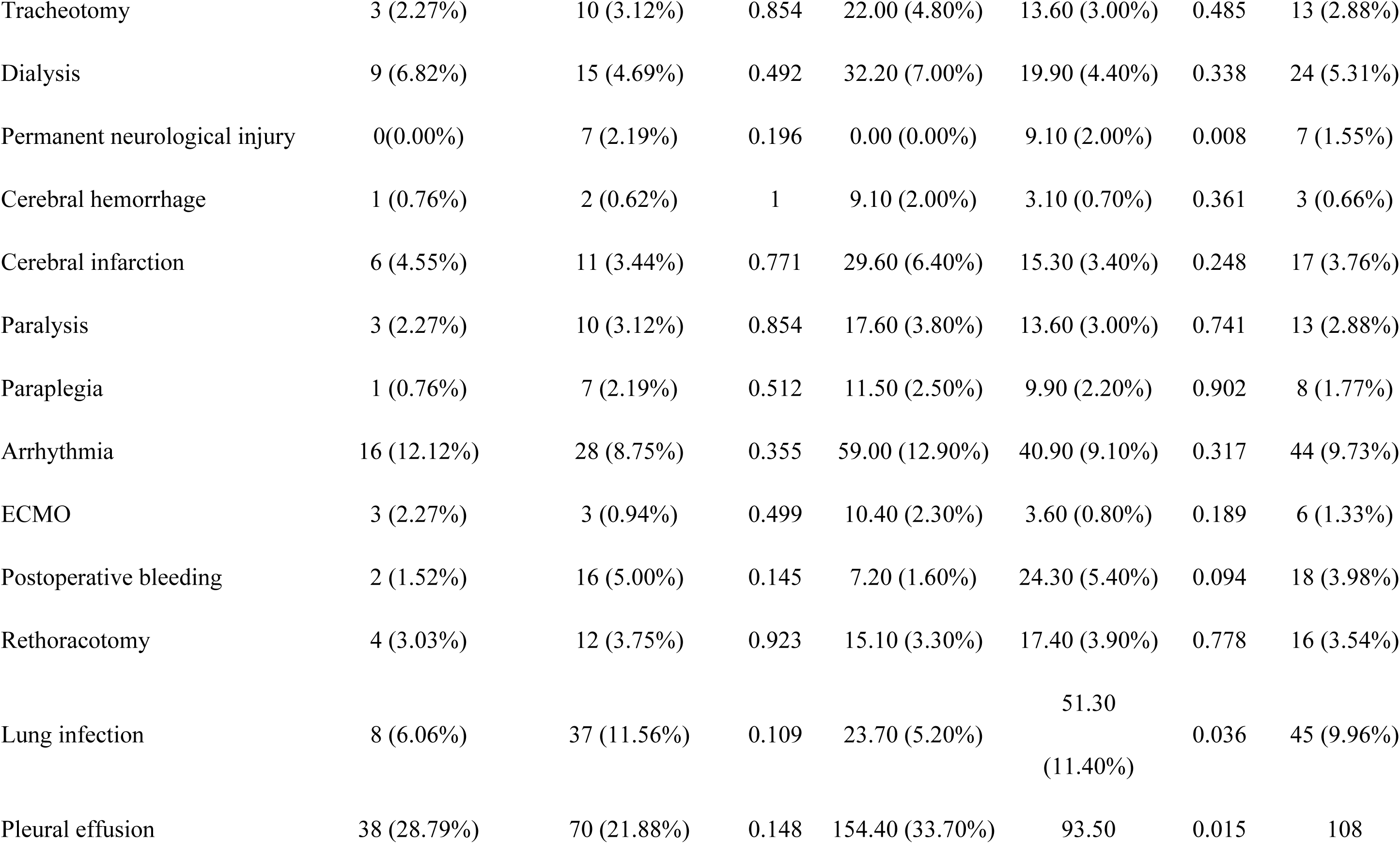
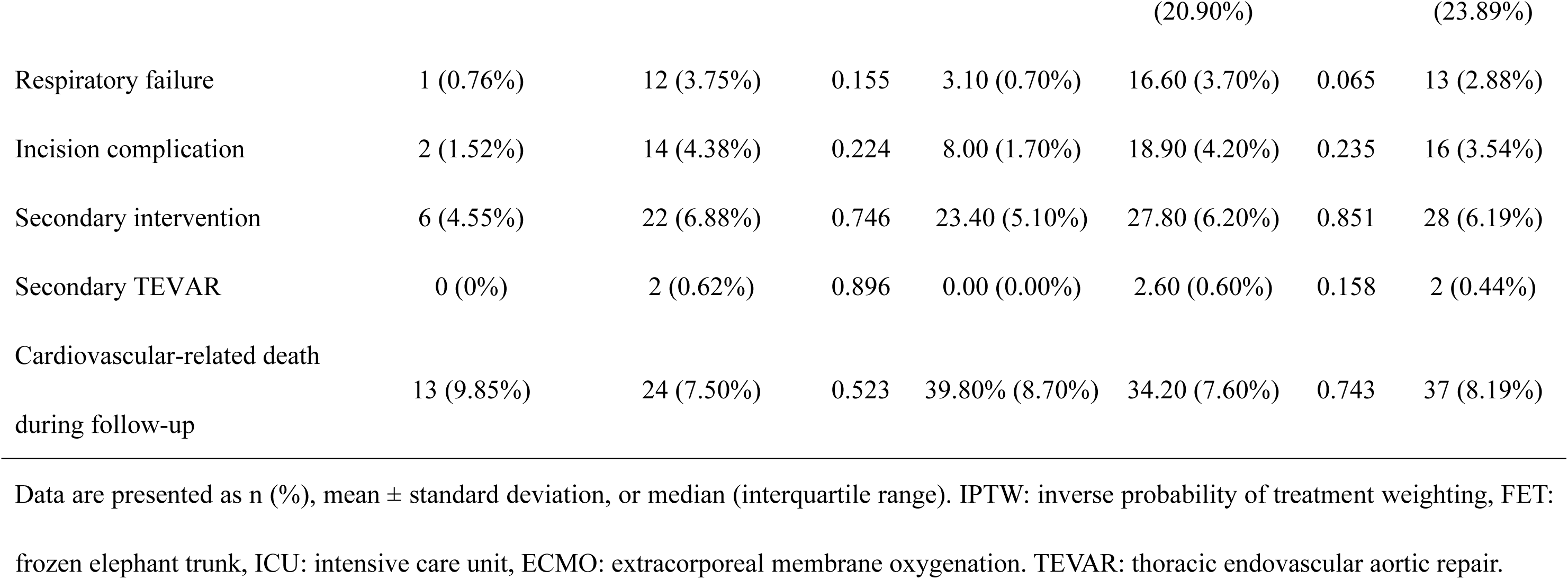
Postoperative data of the total cohort and IPTW cohort.

### LONG-TERM SURVIVAL AND SECONDARY INTERVENTIONS

The median follow-up duration was 3.31 years (IQR 1.82-6.22), with a maximum of 9.24 years. In the unweighted analysis, Kaplan–Meier survival estimates at 1, 3, 5, and 7 years were 93.1% (95% CI 88.8%-97.5%), 87.3% (95% CI 80.5%-94.7%), 84.9% (95% CI 77.0%-93.6%), and 72.3% (95% CI 58.8%-89%) in the non-FET group and 89.0% (95% CI 85.7%-92.5%), 86.2% (95% CI 82.5%-90.1%), 82.5% (95% CI 78.1%-87.2%), and 80.8% (95% CI 75.8%-86.0%) in the FET group, respectively (log-rank P = 0.704; **Figure 2**). After IPTW adjustment, there was no significant difference in the survival rates between the two groups (log-rank P = 0.469, **Figure 2**). Secondary interventions occurred in 6.88% of FET patients and 4.55% of non-FET patients (P = 0.746).

**Figure 2.**
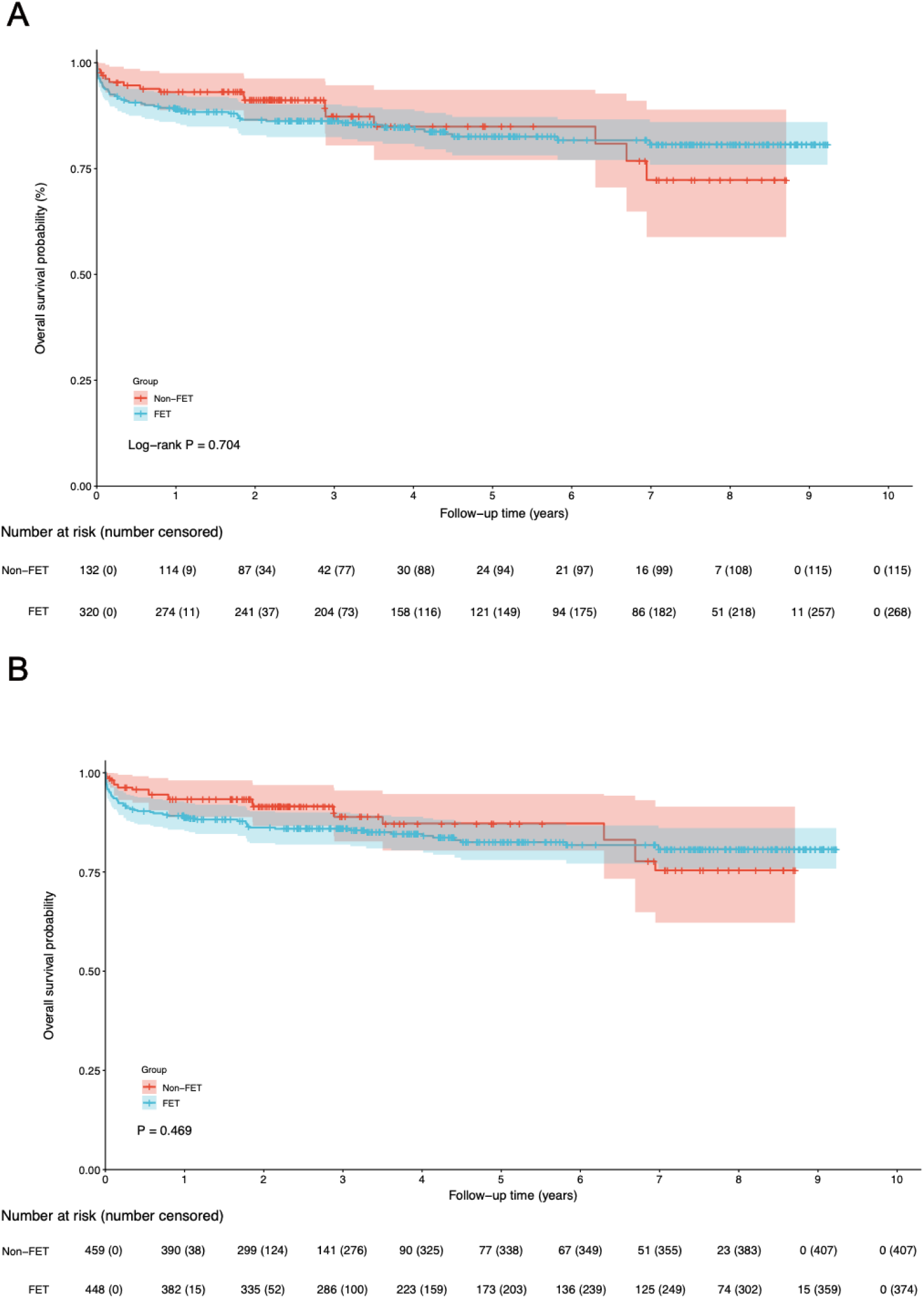
Kaplan–Meier curve for survival before and after IPTW. A) Before IPTW, the survival rates at 1, 3, 5, and 7 years were 93.1% (95% CI 88.8%-97.5%), 87.3% (95% CI 80.5%-94.7%), 84.9% (95% CI 77.0%-93.6%), and 72.3% (95% CI 58.8%-89%) in the non-FET group, and 89.0% (95% CI 85.7%-92.5%), 86.2% (95% CI 82.5%-90.1%), 82.5% (95% CI 78.1%-87.2%), and 80.8% (95% CI 75.8%-86.0%) in the FET group. (log-rank P = 0.705) B) After IPTW, the survival rates at 1, 3, 5, and 7 years were 92.9% (95% CI 88.7%-97.2%), 88.5% (95% CI 82.4%-94.9%), 85.3% (95% CI 79.1%-91.6%), and 74.6% (95% CI 61.7%-87.9%) in the non-FET group, and 89.2% (95% CI 85.8%-92.7%), 85.1% (95% CI 81.0%-89.5%), 81.9% (95% CI 76.7%-86.9%), and 80.2% (95% CI 74.6%-85.6%) in the FET group. (log-rank P = 0.713) IPTW, inverse probability of treatment weighting, FET: frozen elephant trunk.

Secondary TEVAR was rare, performed in only 2 patients (0.62%) in the FET group and none in the non-FET group (P = 0.896). Cardiovascular mortality during follow-up was 7.50% (n = 24) in the FET group versus 9.85% (n = 13) in the non-FET group, with no significant difference (P = 0.523). After IPTW adjustment, these findings remained unchanged (**Table 3**).

Landmark analysis in the IPTW-weighted cohorts revealed no difference in all-cause mortality within the first six years (14.9% vs 18.4%, P = 0.201). Beyond six years, mortality was significantly lower in the FET group (1.8% vs 12.5%, P = 0.007; **Figure 3**).

**Figure 3.**
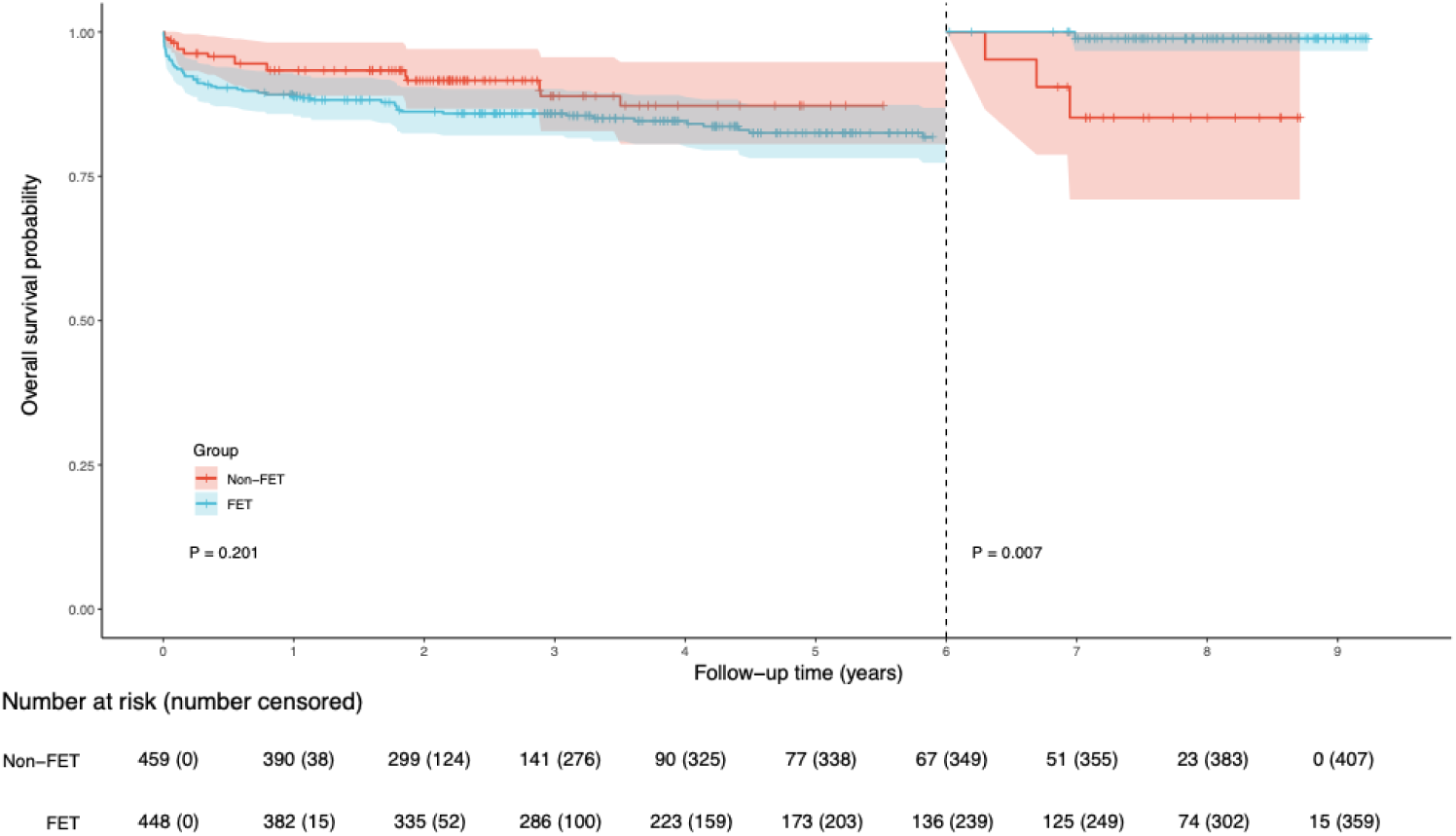
Landmark analysis Kaplan–Meier curve for survival for all patients in a landmark analysis between 6 years and 9 years after IPTW. IPTW, inverse probability of treatment weighting, FET: frozen elephant trunk.

## DISCUSSION

This multicenter study represents one of the largest cohorts analyzing outcomes of subacute and chronic DeBakey type I aortic dissection. Using IPTW and landmark analysis, we demonstrated that the TAR+FET approach was associated with increased early mortality but comparable midterm outcomes and a potential late survival advantage beyond 6 years.

Because of high mortality or prompt surgical intervention, few patients with acute DeBakey type I aortic dissection progress to the subacute/chronic phase^3^. Surgical guidelines for the treatment of subacute/chronic DeBakey type I aortic dissection are limited because of the paucity of data^3^. Concerns about increased operative mortality due to extensive distal repairs have led to the widespread use of a conservative tear-oriented approach, limited to non-FET approach such as ascending aortic (or hemi-arch) replacement, which is considered safe and results in favorable surgical outcomes^6,12^. Although the conservative tear-oriented approach eliminates the risk of cardiac complications and pericardial tamponade, it leaves the distal aortic arch and downstream aorta in a state of dissection, which may lead to postoperative organ malperfusion and a high probability of late intervention^13^. Therefore, this approach is detrimental to long-term survival and increases the likelihood of secondary interventions in the distal aorta, particularly in young patients with aortic dissection.

To address the above-mentioned issues, the FET technique has become an effective option for treating aortic disease when the aortic arch is involved^7^. Existing studies have reported that the FET technique has demonstrated favorable outcomes in the treatment of acute aortic dissection. Specifically, the FET technique facilitates aortic remodeling by promoting false lumen thrombosis not only at the site of stent implantation but also in the distal descending aorta^14^. However, in subacute/chronic aortic dissection, the intima may exhibit hyperplastic or burdened by atherosclerotic plaque, and the adventitia and media may demonstrate increased fibrosis^15^, resulting in a stiffer vessel wall that potentially compromises the efficacy of the FET. Although one theoretical advantage of FET is prevention of future distal reintervention, we did not observe a significant reduction in secondary procedures. This could be related to the relatively short median follow-up and the low absolute rate of late aortic events. Likewise, detailed imaging data on false lumen thrombosis were unavailable for all centers, precluding quantitative analysis of distal remodeling. Future prospective studies should incorporate standardized imaging follow-up to address this question.

The use of the FET technique should be determined based on the specific characteristics of the patient’s aortic pathology. When the arterial supply to organs or the spinal cord originates from the false lumen, thrombosis in the false lumen caused by stent compression can be catastrophic^16^. Weiss et al.^17^ reported their experience using the frozen elephant trunk for simultaneous repair of the aortic arch and descending aorta. The authors defined complete thrombosis of the false lumen as a successful outcome, regardless of the origin of the visceral arteries. However, they encountered malperfusion-related complications, such as spinal cord injury, intestinal ischemia, and permanent dialysis, which occurred exclusively in patients with chronic aortic dissection. Nevertheless, our results show that the postoperative paraplegia frequency among FET patients was 2.19%, and the postoperative dialysis frequency was 4.69%, with no significant difference compared to the non-FET group. For patients with visceral arteries originating from different lumens, we also perform thorough preoperative assessments to mitigate the risk of organ malperfusion caused by false lumen thrombosis. In certain cases, the use of FET may be withheld for such patients.

In terms of baseline characteristics and surgical differences, patients in the FET group had smaller aortic sinus and maximal ascending aorta diameters and underwent fewer root replacements and concomitant valvular procedures. This suggests that patients with significant aortic root dilatation (e.g., diameter >45 mm), severe aortic regurgitation, or organic mitral/tricuspid valve lesions are at higher surgical risk, prompting surgeons to opt for a more conservative non-FET approach to reduce surgical risk and the use of deep hypothermic circulatory arrest (DHCA). In contrast, patients with limited proximal aortic involvement—such as preserved root structure without the need for full root replacement—and good overall cardiac functional reserve are more likely to tolerate the additional steps associated with FET, including stent graft implantation and aortic arch reconstruction. In other words, FET appears to be more suitable for patients with relatively low preoperative surgical risk, allowing them to withstand the procedural complexities and risks associated with the FET technique. This selection bias may explain the slightly higher operative risk but better late stability in the FET group. Overall, with comprehensive preoperative evaluation, the FET technique is safe for subacute or chronic DeBakey type I aortic dissection.

The higher early mortality observed in the FET group likely reflects greater procedural complexity and longer operative times rather than an intrinsic risk of the FET device itself. Similar trends have been observed in large registries of chronic dissection treated with FET. Importantly, the absolute early mortality (6.2%) in our cohort remains lower than that reported in previous chronic dissection series (10-12%)^18,19^. The long-term survival rates for the FET group were 89.0%, 86.2%, 82.5%, and 80.8% at 1, 3, 5, and 7 years, respectively. Compared to reported survival rates for chronic aortic dissection patients treated with FET, which ranged from 66.4% to 76.2% at 5 to 7 years^20–22^, the survival rates in our study were generally higher at each time point. Moreover, our data suggested that there was no significant difference in long-term survival between the FET and non-FET groups before and after IPTW adjustment. Survival curve analysis revealed an intersection of the survival curves for the two groups at 6 years. Landmark analysis showed that TAR+FET may confer potential long-term survival benefit compared with non-FET repair after 6 years. In summary, the present analysis indicates that TAR+FET may be a feasible option for the management of subacute/chronic DeBakey type I aortic dissection.

Schrestha and colleagues^23^ reported that the most common postoperative complications following FET are stroke, paraplegia, and acute kidney injury requiring dialysis. Among these, paraplegia due to spinal cord injury (SCI) is widely recognized as the most critical concern associated with FET. This complication is typically attributed to excessive distal extension of the stent graft and prolonged DHCA. In a meta-analysis, Tian et al. found that 5.1% of patients (range 0-24%) who underwent FET experienced spinal cord injury^24^. Another meta-analysis conducted by Preventza et al.^25^ reported a comparable overall SCI rate of 4.7%. However, we did not find an increase in major adverse events in patients with subacute/chronic DeBakey type I aortic dissection when comparing FET with non-FET. In the FET group, 4.06% (n = 13) of patients had postoperative stroke, 2.19% (n = 7) developed paraplegia due to SCI, and 4.69% (n = 15) required dialysis for kidney injury. In our center, we aimed to reduce the risk of SCI by controlling the depth of the FET distal landing zone (not reach to deeper than the Th7)^11,26^, and using prophylactic cerebrospinal fluid drainage in high-risk patients ^27^. To prevent malperfusion, especially in obese patients or those with extensive dissection involving multiple branch vessels or significant true lumen narrowing, we typically avoid single-site cannulation and instead adopt a DAC strategy. Single-site cannulation—particularly via the axillary artery—may prolong cooling time and result in uneven temperature distribution in overweight patients. Moreover, in cases of true lumen compression or branch vessel malperfusion, it may fail to ensure adequate systemic perfusion.

## LIMITATION

This study has several limitations. First, the retrospective design of the study is a primary limitation. Although IPTW was used to adjust for confounding factors, it is still challenging to fully eliminate selection bias and information bias. Second, given the median follow-up time of only 3.31 years in this study, the result that the FET group has superior long-term survival after 6 years, derived from data extrapolation, should be interpreted cautiously. Thus, it can only be inferred that FET may offer potential late clinical benefits. Prospective studies with longer follow-up periods are needed to validate if the findings are robust. Third, variations in surgical expertise across centers may influence perioperative outcomes. Finally, the absence of complete imaging data on distal remodeling and false lumen thrombosis limited our ability to assess anatomical benefits of the FET procedure. Future large-scale prospective randomized controlled trials are needed to further validate the findings of this study.

## CONCLUSION

In patients with subacute or chronic DeBakey type I dissection, TAR+FET is a feasible surgical method associated with increased early mortality, but potential late survival benefit compared with non-FET approaches. These findings warrant confirmation in prospective studies with extended follow-up.

## Data Availability

The data that support the findings of this study are not publicly available due to restrictions containing information that could compromise the privacy of research participants. De-identified data may be made available from the corresponding author upon reasonable request and with approval from the relevant institutional review board(s)/ethics committee(s) and data-sharing agreements.

## ACKNOWLEDGEMENTS

The authors would like to thank AiMi (www.aimieditor.com) for providing linguistic assistance.

## DISCLOSURE STATEMENT

The authors declare no potential conflicts of interest with respect to the research, authorship, and/or publication of this article.

**Abbreviations**
AKI: Acute kidney injury
BMI: Body mass index
CABG: Coronary artery bypass graft
CPB: Cardiopulmonary bypass
DAC: Dual arterial cannulation
FET: Frozen elephant trunk
IPTW: Inverse probability of treatment weighting
LVEF: Left ventricular ejection fraction
MVR: Mitral valve replacement
SCP: Selective cerebral perfusion
TAR: Total arch replacement
TEVAR: Thoracic endovascular aortic repair
TVr: Tricuspid valve repair

## REFERENCES

1. Takagi S, Goto Y, Yanagisawa J, Ogihara Y, Okawa Y. Strategy for acute DeBakey type I aortic dissection considering midterm results: a retrospective cohort study comparing ascending aortic replacement and total arch replacement with frozen elephant trunk technique. J Cardiothorac Surg. 2024;19(1):15. doi:10.1186/s13019-024-02484-6

2. Czerny M, Grabenwöger M, Berger T, et al. EACTS/STS Guidelines for Diagnosing and Treating Acute and Chronic Syndromes of the Aortic Organ. The Annals of Thoracic Surgery. 2024;118(1):5–115. doi:10.1016/j.athoracsur.2024.01.021

3. Wu J, Xie E, Qiu J, et al. Subacute/chronic type A aortic dissection: a retrospective cohort study. European Journal of Cardio-Thoracic Surgery. Published online July 16, 2019:ezz209. doi:10.1093/ejcts/ezz209

4. Raffa GM, Pilato M, Armaro A, Ruperto C, Gentile G, Follis F. Chronic Stanford type A aortic dissection: Journal of Cardiovascular Medicine. 2016;17:e138–e140. doi:10.2459/JCM.0000000000000175

5. Hynes C, Greenberg M, Sarin S, Trachiotis G. Chronic Type A Aortic Dissection: Two Cases and a Review of Current Management Strategies. Aorta. 2016;04(01):16–21. doi:10.12945/j.aorta.2015.15.016

6. Rylski B, Milewski RK, Bavaria JE, et al. Outcomes of Surgery for Chronic Type A Aortic Dissection. The Annals of Thoracic Surgery. 2015;99(1):88–93. doi:10.1016/j.athoracsur.2014.07.032

7. Chivasso P, Mastrogiovanni G, Miele M, et al. Frozen Elephant Trunk Technique in Acute Type A Aortic Dissection: Is It for All? Medicina. 2021;57(9):894. doi:10.3390/medicina57090894

8. Sun L, Qi R, Zhu J, Liu Y, Zheng J. Total Arch Replacement Combined With Stented Elephant Trunk Implantation: A New “Standard” Therapy for Type A Dissection Involving Repair of the Aortic Arch? Circulation. 2011;123(9):971–978. doi:10.1161/CIRCULATIONAHA.110.015081

9. Sun LZ, Qi RD, Chang Q, et al. Is total arch replacement combined with stented elephant trunk implantation justified for patients with chronic Stanford type A aortic dissection? The Journal of Thoracic and Cardiovascular Surgery. 2009;138(4):892–896. doi:10.1016/j.jtcvs.2009.02.041

10. Liang S, Liu Y, Zhang B, et al. Cannulation strategy in frozen elephant trunk for type A aortic dissection: double arterial cannulation approach. European Journal of Cardio-Thoracic Surgery. 2022;62(3):ezac165. doi:10.1093/ejcts/ezac165

11. Iino K, Takago S, Saito N, et al. Total arch replacement and frozen elephant trunk for acute type A aortic dissection. The Journal of Thoracic and Cardiovascular Surgery. 2022;164(5):1400–1409.e3. doi:10.1016/j.jtcvs.2020.10.135

12. Silaschi M, Byrne J, Wendler O. Aortic dissection: medical, interventional and surgical management. Heart. 2017;103(1):78–87. doi:10.1136/heartjnl-2015-308284

13. Evangelista A, Salas A, Ribera A, et al. Long-Term Outcome of Aortic Dissection With Patent False Lumen: Predictive Role of Entry Tear Size and Location. Circulation. 2012;125(25):3133–3141. doi:10.1161/CIRCULATIONAHA.111.090266

14. Papakonstantinou NA, Martinez-Lopez D, Chung JCY. The frozen elephant trunk: seeking a more definitive treatment for acute type A aortic dissection. European Journal of Cardio-Thoracic Surgery. 2024;65(5):ezae176. doi:10.1093/ejcts/ezae176

15. Peterss S, Mansour AM, Ross JA, et al. Changing Pathology of the Thoracic Aorta From Acute to Chronic Dissection. Journal of the American College of Cardiology. 2016;68(10):1054–1065. doi:10.1016/j.jacc.2016.05.091

16. Urbanski PP, Bougioukakis P, Deja MA, et al. Open aortic arch surgery in chronic dissection with visceral arteries originating from different lumens. Eur J Cardiothorac Surg. 2016;49(5):1382–1390. doi:10.1093/ejcts/ezv386

17. Weiss G, Tsagakis K, Jakob H, et al. The frozen elephant trunk technique for the treatment of complicated type B aortic dissection with involvement of the aortic arch: multicentre early experience†. European Journal of Cardio-Thoracic Surgery. 2015;47(1):106–114. doi:10.1093/ejcts/ezu067

18. Panfilov DS, Kozlov BN. Mid-term Outcomes of Frozen Elephant Trunk for Chronic Aortic Dissection. Canadian Journal of Cardiology. 2025;41(5):989–995. doi:10.1016/j.cjca.2025.01.003

19. Pacini D, Tsagakis K, Jakob H, et al. The Frozen Elephant Trunk for the Treatment of Chronic Dissection of the Thoracic Aorta: A Multicenter Experience. The Annals of Thoracic Surgery. 2011;92(5):1663–1670. doi:10.1016/j.athoracsur.2011.06.027

20. Arnold Z, Geisler D, Aschacher T, et al. Long-Term Results with 187 Frozen Elephant Trunk Procedures. JCM. 2023;12(12):4143. doi:10.3390/jcm12124143

21. Yamane Y, Katayama K, Furukawa T, et al. Mid-Term Results of Frozen Elephant Trunk Technique for Chronic Aortic Dissection. Annals of Vascular Diseases. 2020;13(2):137–143. doi:10.3400/avd.oa.19-00131

22. Luo C, Qi R, Zhong Y, et al. Early and Long-Term Follow-Up for Chronic Type B and Type Non-A Non-B Aortic Dissection Using the Frozen Elephant Trunk Technique. Front Cardiovasc Med. 2021;8:714638. doi:10.3389/fcvm.2021.714638

23. Shrestha M, Bachet J, Bavaria J, et al. Current status and recommendations for use of the frozen elephant trunk technique: a position paper by the Vascular Domain of EACTS. Eur J Cardiothorac Surg. 2015;47(5):759–769. doi:10.1093/ejcts/ezv085

24. Tian DH, Wan B, Eusanio MD, Black D, Yan TD. A systematic review and meta-analysis on the safety and efficacy of the frozen elephant trunk technique in aortic arch surgery. Annals of cardiothoracic surgery. 2013;2(5).

25. Preventza O, Liao JL, Olive JK, et al. Neurologic complications after the frozen elephant trunk procedure: A meta-analysis of more than 3000 patients. The Journal of Thoracic and Cardiovascular Surgery. 2020;160(1):20–33.e4. doi:10.1016/j.jtcvs.2019.10.031

26. Flores J, Kunihara T, Shiiya N, Yoshimoto K, Matsuzaki K, Yasuda K. Extensive deployment of the stented elephant trunk is associated with an increased risk of spinal cord injury. The Journal of Thoracic and Cardiovascular Surgery. 2006;131(2):336–342. doi:10.1016/j.jtcvs.2005.09.050

27. Spratt JR, Walker KL, Neal D, et al. Rescue therapy for symptomatic spinal cord ischemia after thoracic endovascular aortic repair. The Journal of Thoracic and Cardiovascular Surgery. 2024;168(1):15–25.e11. doi:10.1016/j.jtcvs.2022.10.045

